# Pulmonary post-mortem findings in a large series of COVID-19 cases from Northern Italy

**DOI:** 10.1101/2020.04.19.20054262

**Authors:** Luca Carsana, Aurelio Sonzogni, Ahmed Nasr, Roberta Simona Rossi, Alessandro Pellegrinelli, Pietro Zerbi, Roberto Rech, Riccardo Colombo, Spinello Antinori, Mario Corbellino, Massimo Galli, Emanuele Catena, Antonella Tosoni, Andrea Gianatti, Manuela Nebuloni

## Abstract

**Importance:** The analysis of lung tissues of patients with COVID-19 may help understand pathogenesis and clinical outcomes in this life-threatening respiratory illness.

**Objective:** To determine the histological patterns in lung tissue of patients with severe COVID-19.

**Design and participants:** Lungs tissues of 38 cases who died for COVID-19 in two hospital of Northern Italy were systematically analysed. Hematoxylin-eosin staining, immunohistochemistry for the inflammatory infiltrate and cellular components, electron microscopy were performed.

**Results:** The features of the exudative and proliferative phases of Diffuse Alveolar Disease (DAD) were found: capillary congestion, necrosis of pneumocytes, hyaline membrane, interstitial oedema, pneumocyte hyperplasia and reactive atypia, platelet-fibrin thrombi. The inflammatory infiltrate was composed by macrophages in alveolar lumens and lymphocytes mainly in the interstice. Electron microscopy revealed viral particles in the cytoplasm of pneumocytes.

**Conclusions and relevance:** The predominant pattern of lung lesions in COVID-19 patients is DAD, as described for the other two coronavirus that infect humans, SARS-CoV and MERS-CoV. Hyaline membrane formation and pneumocyte atypical hyperplasia are frequently found. The main relevant finding is the presence of platelet-fibrin thrombi in small arterial vessels; this important observation fits into the clinical context of coagulopathy which dominates in these patients and which is one of the main targets of therapy.

## INTRODUCTION

Since December 2019, an outbreak caused by a new coronavirus infection (severe acute respiratory syndrome coronavirus 2, SARS-CoV-2) disease was reported, starting in China but rapidly spreading in many countries all over the world. Italy has been the first country in Europe to be reached by the epidemic and Lombardy was devastated in just one month. L.Sacco Hospital, Milan and Papa Giovanni XXIII, Bergamo have been the first hospitals in this region to manage the epidemic crisis.

The clinical spectrum of SARS-CoV-2 disease (COVID-19) is reported to include mild asymptomatic infection, mild upper respiratory disease with fever and cough, and severe pneumonia that can lead to ARDS in 15% of the hospitalized cases^1^. Although the epidemic outbreak started at the end of the last year, no systematic reviews of the pathological features of COVID-19 lung involvement have been published so far.

We describe the first available large series of lung histopathological findings in patients died from COVID-19 in Northern Italy, with the aim to report the main microscopic pulmonary lesions in SARS-CoV-2 infection and severe respiratory failure.

## METHODS

This study is based on the histological analysis of post-mortem lung tissues from 38 cases who died for COVID-19 between February and March in two referral centres for the management of the COVID-19 outbreak in Northern Italy, Luigi Sacco Hospital, Milan, and Papa Giovanni XXIII, Bergamo (patient permission given by the Ethics Committees of the two hospitals to use personal and sensitive data for scientific research related to the disease was collected).

The autopsies were performed in Airborne Infection Isolation Autopsy Rooms and the personnel used the correct Personal Protection Equipment (PPE), according to “Engineering control and PPE recommendations for autopsies”.

A medium of 7 tissue blocks were taken from each lung (range 5-9), selecting the most representative areas at macroscopic examination. Tissues were fixed in 10% buffered formalin for >48 hours. Three-μm paraffin sections were stained by hematoxylin-eosin. Immunohistochemistry reactions were performed on selected cases (CD45, CD68, CD61, TTF1, p40, Ki67, Masson Trichome) to better characterize inflammatory infiltrate, epithelial cells and fibrosis. Histological evaluation was performed blindly by two pathologists from each hospital, with expertise in the field. Histological features of the cellular and interstitial damage were described and graded by using a semiquantitative scale.

Additional samples from selected cases were fixed in glutaraldehyde for electron microscopy and examined by EM-109 ZEISS and CCD-Megaview G2 (I-TEM imaging platform software).

## RESULTS

Patients were 33 males and five females, average age of 69 years (range 32-86); the time that the patients spent in the Subintensive/Intensive Care Unit ranged from 1 to 23 days (6.87 days). Regarding past comorbidities, data were available in 31/38 patients: 9 diabetes, 18 hypertension, 4 past malignancies, 11 cardiovascular disorders, 3 mild chronic obstructive pulmonary disorders. At the time of hospitalization, all the patients had throat swab sample positive for SARS-CoV-2 infection and had clinical and radiological features of interstitial pneumonia. D-dimer was available in 26/38 patients, with high value in all of them (>10 × the upper reference limit). Patients died after a median time of 16.27 days (range 5-31) from the onset of symptoms.

Macroscopic examination of the lungs revealed heavy, congested and oedematous organs, with spotty involvement. At histological examination, the features of Diffuse Alveolar Disease (DAD) were found, corresponding to those observable in the exudative and early/intermediate proliferative phases of the disease. Both phases often overlapped in the different areas of the lungs, with plurifocal pattern of distribution. The fibrotic phase was rarely observed, possibly due to the short duration of the disease. Moreover, five patients also had bacterial (4) and fungal (1) abscesses.

Table 1 reported all the morphological parameters, with the corresponding semiquantitative grading. Capillary congestion, interstitial oedema, dilated alveolar ducts, hyaline membranes composed of serum proteins and condensed fibrin, loss of pneumocytes were the histological patterns of the exudative phase mostly observed in all the cases. Platelet-fibrin thrombi in small arterial vessels (<1mm in diameter) were found in 33 cases. Moreover, type II pneumocyte hyperplasia showing reactive atypia, myofibroblast proliferation, alveolar granulation tissue and obliterating fibrosis were present in half of the patients but were focal. Microcystic honeycombing and mural fibrosis were occasionally present. The main histological features are reported in Figure 1.

**Table 1.** Histological data

|  |  | Absent | Rare | Focal | Present | Plurifocal | Diffuse |
| --- | --- | --- | --- | --- | --- | --- | --- |
| **Exudative phase** | Capillary congestion | 0 | 0 | 0 | 24 | 1 | 13 |
|  | Interstitial and intraalveolar edema | 1 | 0 | 19 | 10 | 5 | 3 |
|  | Alveolar proteinosis | 10 | 0 | 25 | 1 | 2 | 0 |
|  | Alveolar hemorrhage | 5 | 1 | 20 | 8 | 2 | 2 |
|  | Hyaline membranes | 5 | 1 | 19 | 5 | 3 | 5 |
|  | Dilated alveolar ducts plus collapsed alveoli | 2 | 0 | 16 | 18 | 2 | 0 |
|  | Collapsed alveoli | 9 | 1 | 7 | 21 | 0 | 0 |
|  | Increased megakaryocytes | 5 | 0 | 25 | 4 | 3 | 1 |
|  | Granulocytes | 6 | 1 | 14 | 14 | 2 | 1 |
|  | Loss of pneumocytes | 0 | 0 | 11 | 20 | 3 | 4 |
|  | Fibrin thrombi | 5 | 0 | 16 | 4 | 13 | 0 |
|  | Localized DIC | 38 | 0 | 0 | 0 | 0 | 0 |
| **Proliferative phase** | Type 2 pneumocyte hyperplasia | 0 | 0 | 14 | 9 | 8 | 7 |
|  | Squamous metaplasia with atypia | 17 | 1 | 12 | 7 | 1 | 0 |
|  | Interstitial myofibroblast reaction | 13 | 0 | 18 | 6 | 1 | 0 |
|  | Alveolar granulation tissue | 17 | 0 | 13 | 3 | 4 | 1 |
|  | Alveolar occludent fibrosis (complete) | 26 | 1 | 6 | 3 | 1 | 1 |
|  | Alveolar occludent fibrosis ("ring" fibrosis) | 23 | 0 | 10 | 3 | 1 | 1 |
|  | Septal collagen deposition | 23 | 0 | 13 | 1 | 1 | 0 |
|  | Alveolar duct fibrosis | 26 | 0 | 12 | 0 | 0 | 0 |
|  | Alveolar buds | 27 | 1 | 7 | 3 | 0 | 0 |
|  | Capillaries proliferation | 20 | 0 | 14 | 3 | 0 | 1 |
|  | Bronchiolitis obliterans | 38 | 0 | 0 | 0 | 0 | 0 |
|  | Organized alveoli plus dilated alveolar ducts | 29 | 0 | 6 | 3 | 0 | 0 |
| **Fibrotic phase** | Pleural involvement | 38 | 0 | 0 | 0 | 0 | 0 |
|  | Mural fibrosis | 14 | 0 | 12 | 10 | 1 | 1 |
|  | Scars | 38 | 0 | 0 | 0 | 0 | 0 |
|  | Fibrous microcysts | 37 | 0 | 1 | 0 | 0 | 0 |
|  | Microcystic honeycombing | 23 | 0 | 9 | 6 | 0 | 0 |
|  | Arterial hypermuscularization | 34 | 1 | 2 | 1 | 0 | 0 |
| **Other associated lesions** | Interstitial inflammatory infiltrate | 7 | 0 | 5 | 12 | 10 | 4 |
|  | Alveolar inflammatory infiltrate (macrophages) | 14 | 1 | 13 | 8 | 0 | 2 |
|  | Alveolar multinucleated giant cells | 19 | 6 | 9 | 1 | 1 | 2 |

Absent: 0; rare: <5% of the tissue; focal: 5-25% of the tissue; present: 25-50% of the tissue; plurifocal: 50-75% of the tissue; diffuse: >75% of the tissue.

**Figure 1.**
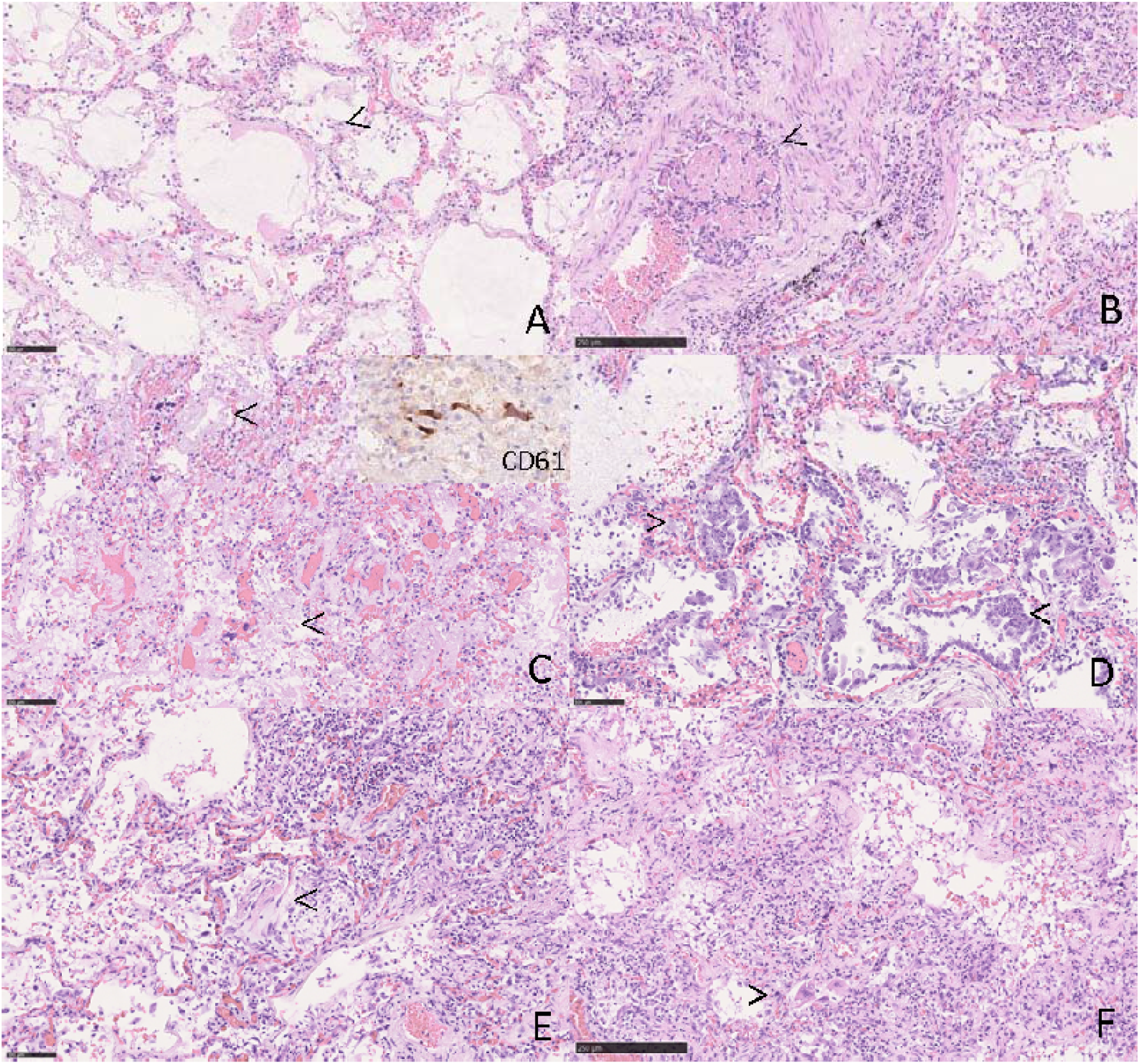
Lung parenchima with diffuse alveolar damage: Panel A: exudative phase with jaline membranes (arrow); Panel B: organising microthrombus (arrow); Panel C: entrapped megakariocytes in alveolar capillaries (arrow), highlightened by CD61 (inset); Panel D: early proliferative phase with many hyperplastic, seldom atypical, type II pneumocytes (arrows); Panel E: intermediate phase with luminal organizing fibrosis (arrow); Panel F: advanced proliferative phase, with interstitial myofibroblastic reaction and residual scattered hyperplastic type II pneumocytes (arrow). Hematoxylin-Eosin, OM 10x

Inflammatory component was represented by a few CD45 positive lymphocytes located in the interstitial space; a large number of CD68 positive macrophages were mainly localized in the alveolar lumens (Figure 2, panels A and B). Immunohistochemistry with anti CD61 antibody identified increased number of megakaryocytes in lung capillaries (Figure 1, panel C, inset).

**Figure 2.**
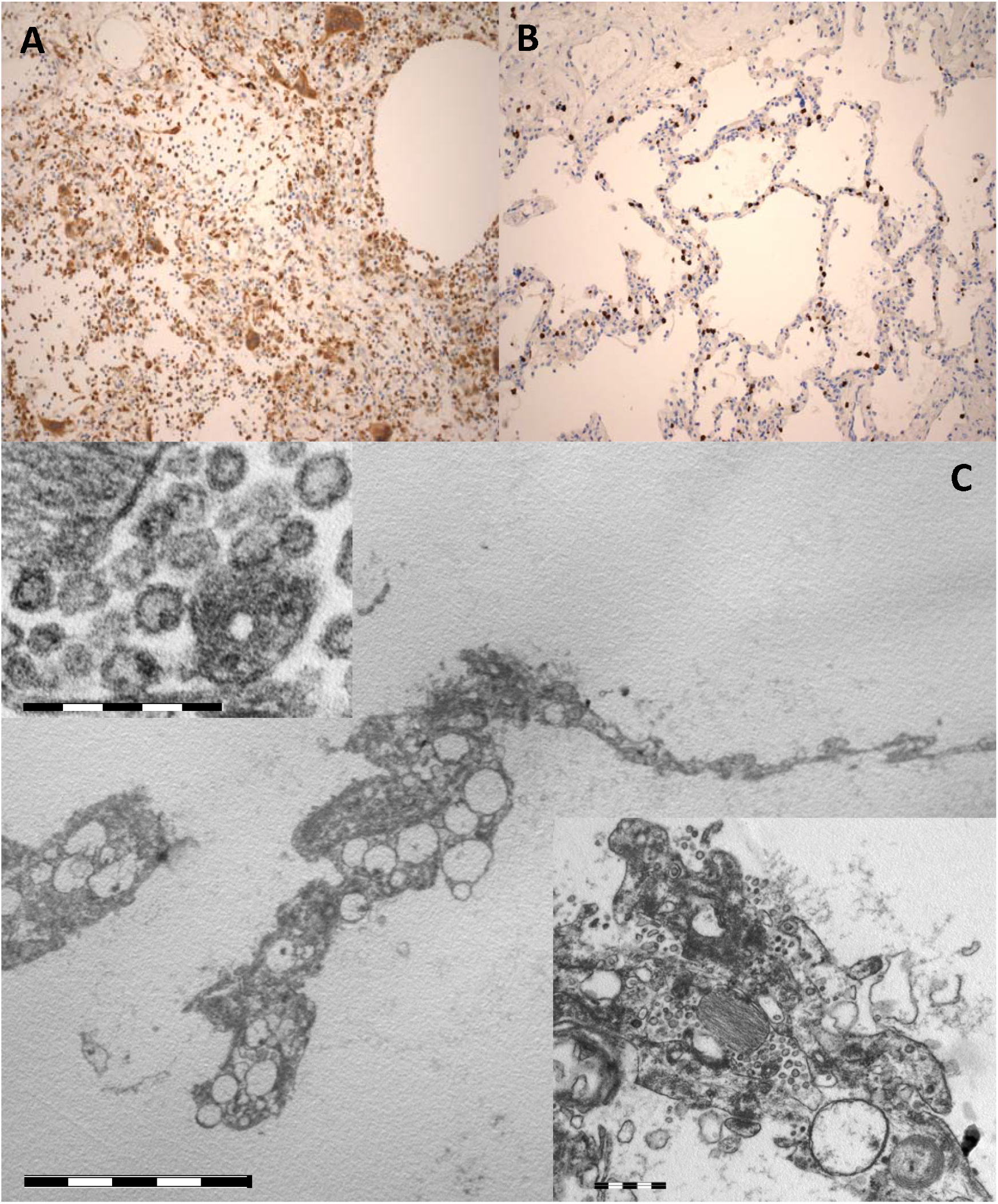
Panel A and B: immunohistochemistry with antibody to CD68 (A) shows highly predominant component of macrophages, focally with multinucleated cells feature. Immunohistochemistry with antibody to CD45 shows a few lymphocytes in interstitial septa (CD68 and CD45 brown immunostaining, Hematoxylin counterstaining, OM x20). Panel C: pneumocyte free in alveolar space and coronavirus viral particles within cytoplasmic vacuoles (inset right down). Virions had an average diameter of 82 nm and viral projection about 13nm in length (inset left up, OMx85000). Bars: 5μm, 500nm, 200nm

Ultrastructural examination revealed viral particles, with morphology typical of the family of Coronaviridae and localized along plasmalemmal membranes and within cytoplasmic vacuoles of pneumocytes (Figure 2, panel C). Virions had an average diameter of 82nm and viral projection about 13nm in length.

## DISCUSSION

Here we report the largest series of COVID-19 autopsies focusing on pulmonary lesions, from patients died in the Northern Italy. In all examinations diffuse pattern of exudative and early proliferative phases of DAD were found, while the fibrotic phase was rare. The peculiar histopathological findings were atypical pneumocytes (reactive atypia) and diffuse thrombosis of the peripheral small vessels.

SARS-CoV-2 is the seventh member of the coronavirus family that causes disease in humans. Two other members of this family are SARS-CoV and MERS-CoV. The three coronaviruses show many similarities in the clinical presentation. SARS-CoV and MERS-CoV can cause acute DAD, associated with pneumocyte hyperplasia and interstitial pneumonia^2, 3^. Both viruses have been described in pneumocytes, macrophages, lung interstitial cells by electron microscopy, immunohistochemistry and ISH^2-6^.

Despite the importance of lung involvement in COVID-19 patients, only limited data are available concerning lung pathology. A recent paper from China described the histological lesions in a patient died of COVID-19; desquamation of pneumocytes, DAD and oedema were found^7^. Other authors described pulmonary pathology of early-phase COVID-19 in two patients with lung carcinoma; both patients exhibited signs of the exudative phase of DAD^8^. Luo et al. performed lung organ dissection and described the pathological changes of one COVID-19 critical patients in the whole organs; features of DAD and vascular occlusion were found (personal observations).

In our study, fibrin thrombi of small arterial vessels (diameter < 1mm) were observed in 33 /38 patients, half of them with >25% of tissue involvement and associated with high levels of D-dimer in blood. These findings might explain the severe hypoxemia which characterizes the clinical feature of ARDS in SARS-CoV-2 patients. Our data strongly support the hypothesis proposed by recent clinical studies, that COVID-19 is complicated or anyway strictly related to coagulopathy and thrombosis; moreover detection of D-dimer values >1 μg/ml have been associated with fatal outcome of COVID-19. For these reasons, the use of anticoagulants has been recently suggested as potentially beneficial in patients with severe COVID-19, albeit its efficacy and safety have not been demonstrated^9^.

Finally, the search for viral particles was carried out in a subset of patients and highlighted the presence of rare virions in the cytoplasm of pneumocytes. Despite the low number of cases, these findings may suggest that the virus remains in lung tissue for many days, even if in small quantities, possibly being the trigger of the mechanism that leads to and feeds lung damage. Further histological and molecular analyses and extension of the series are ongoing to better define cellular and tissue distribution of viruses and organ inflammatory response.

## Data Availability

All data are available by the authors.

